# Left Atrial Calcification After Atrial Fibrillation Ablation Predicts Long-term Risk of Heart Failure Hospitalization

**DOI:** 10.64898/2025.12.01.25341418

**Authors:** Tadanori Nakata, Naoyuki Miwa, Satoshi Hara, Hikaru Shimosato, Mirei Setoguchi, Taiki Ishizawa, Hidenori Hirano, Yoshikazu Sato, Shigeki Kusa, Hitoshi Hachiya, Tetsuo Sasano

**Author notes:** Address for correspondence: Tadanori Nakata, M.D. Department of Cardiology, Tsuchiura Kyodo Hospital 4-1-1 Otsuno, Tsuchiura, Ibaraki 300-0028 Japan. Disclosures: The authors have no relevant disclosure.

## Abstract

**Background:** Left atrial calcification (LAC) after atrial fibrillation (AF) ablation has recently been described, but its clinical significance and determinants remain insufficiently defined.

**Methods:** Among 1,741 patients who underwent first-time radiofrequency AF ablation between 2011 and 2018, 231 patients underwent both pre- and post-ablation chest CT. Patients were classified into an LAC group (n=71) and a non-LAC group (n=160). Baseline characteristics, ablation parameters, and calcification sites were analyzed. Independent predictors of LAC formation were identified using Cox proportional hazards models. Clinical outcomes, including heart failure (HF) hospitalization, stroke, and a composite endpoint of all-cause mortality, HF, stroke, or coronary intervention, were compared during long-term follow-up.

**Results:** Over a mean follow-up of 9.5 ± 2.7 years, LAC was detected in 31% of patients, typically 7.3 ± 3.1 years after ablation. LAC occurred most frequently at the lateral mitral annulus and right pulmonary veins. Independent predictors of LAC formation were non-paroxysmal AF (HR 1.7, 95% CI 1.0–2.8, p=0.04), LA diameter >50 mm (HR 2.7, 95% CI 1.3–5.5, p<0.01), and ablation beyond the pulmonary veins (HR 2.3, 95% CI 1.3–4.2, p=0.04). Kaplan–Meier analysis showed significantly higher HF hospitalization in the LAC group (log-rank p=0.02), while stroke incidence was similar between groups (p=0.50). The composite endpoint occurred more frequently in the LAC group (p=0.05).

**Conclusion:** LAC is a not uncommon late finding after AF ablation, particularly in patients with advanced atrial remodeling or extensive ablation. Its presence is associated with a higher risk of HF hospitalization, underscoring the importance of procedural planning and long-term imaging surveillance in AF ablation patients.

## Introduction

Pulmonary vein isolation is the standard treatment for atrial fibrillation (AF) and has been shown to be more effective than medical therapy in maintaining sinus rhythm^1,2^. Left atrial calcification (LAC) has been described in association with rheumatic fever, mitral valve disease, and organized thrombus^3–5^. More recently, several reports have described the occurrence of LAC following AF ablation^6^. However, its clinical significance remains poorly understood.

We previously encountered a patient in whom the morphology and mobility of post-ablation calcification indicated a high embolic risk, ultimately necessitating surgical removal^7^. This case highlighted that post-ablation calcification in the chronic phase may serve as a potential source of cerebral embolism. Moreover, emerging evidence suggests that LAC after ablation may be associated with subsequent increases in heart failure (HF) hospitalizations^6^.

A prior study demonstrated that LAC can be a prognostic marker of adverse cardiovascular outcomes following AF ablation, but the available data remain limited by sample size, follow-up duration, and incomplete characterization of procedural predictors. To further clarify these issues, we investigated the long-term prognostic significance of LAC after AF ablation in a large single-center cohort, with a focus on both clinical outcomes and procedural factors contributing to LAC formation.

## Methods

### Study population

We retrospectively analyzed 1,741 consecutive patients who underwent a first radiofrequency ablation for AF at Tsuchiura Kyodo Hospital between 2011 and 2018. Among these, 231 patients underwent both pre- and post-ablation CT imaging and were included in the present study. Patients treated with cryoballoon ablation (n=353) and those without CT follow-up ≥1 year after ablation (n=1,388) were excluded (Figure 1). Of the final cohort, 71 patients developed new LAC (Figure 2) and 160 did not. The study was approved by the institutional review board, and all patients provided written informed consent in accordance with the Declaration of Helsinki.

**Figure 1.**
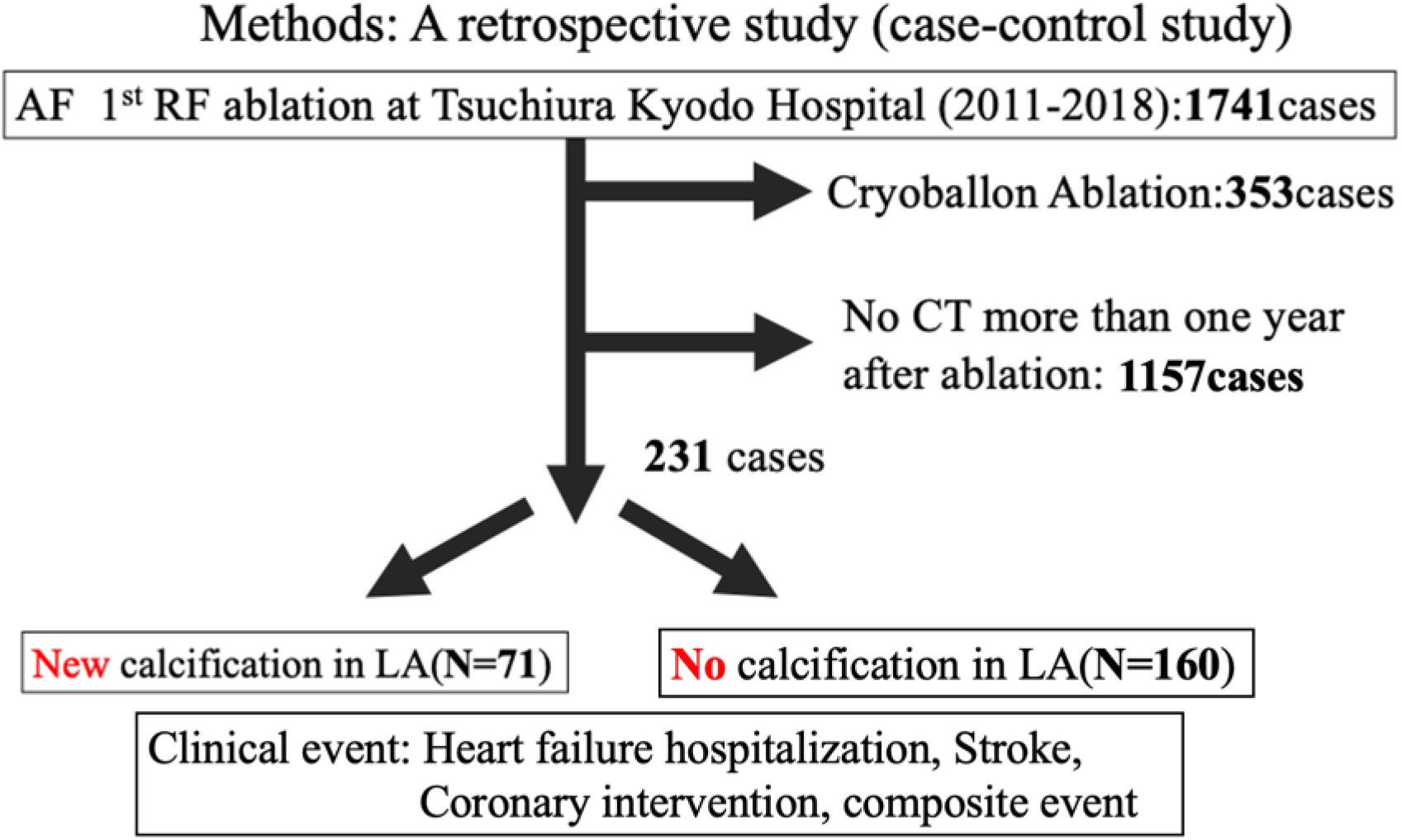
Chronic phase calcification formation in left atrium after RF ablation.

**Figure 2.**
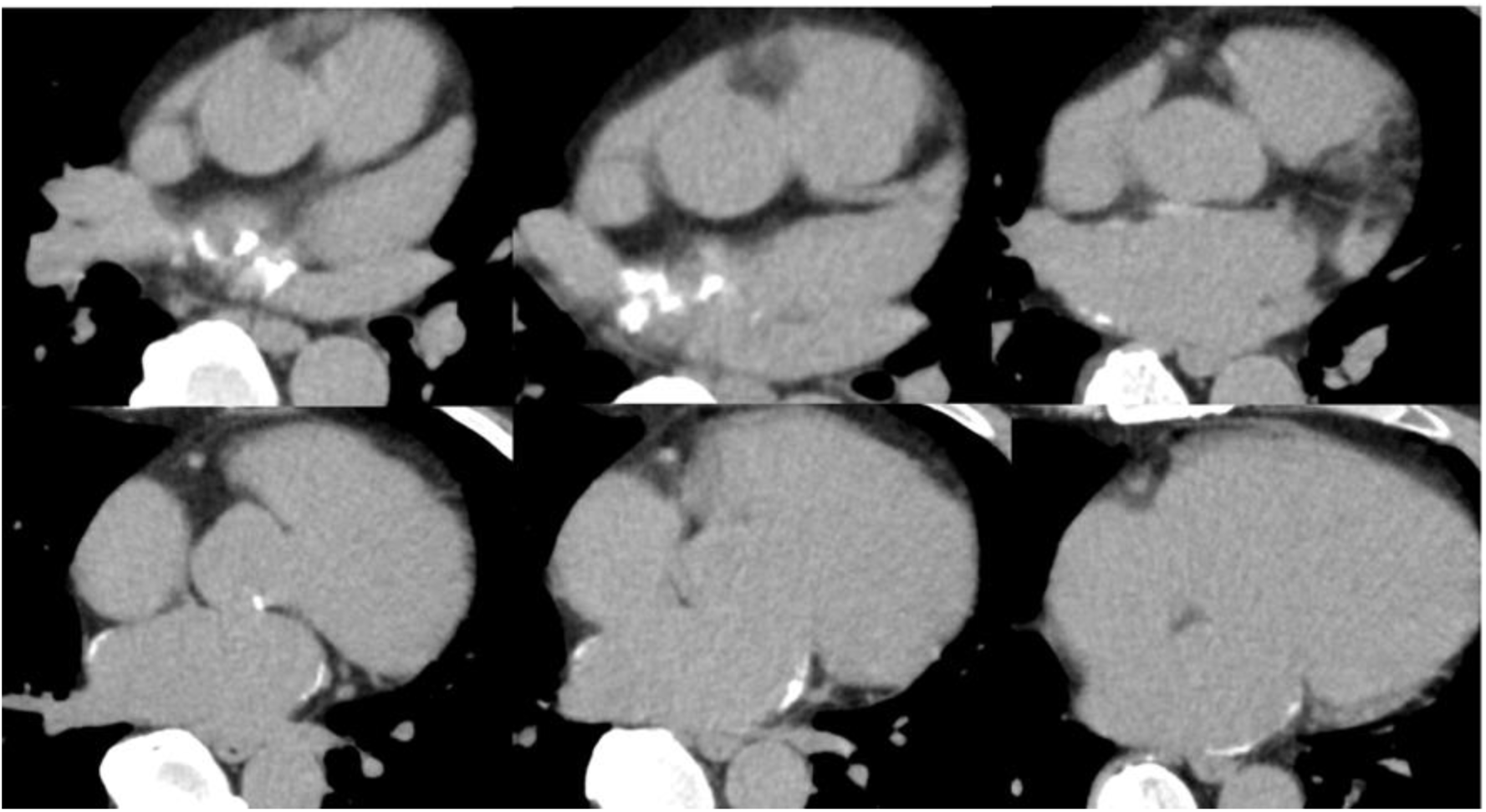
Left Atrial Calcification After Atrial Fibrillation Ablation.

### Ablation procedures

Procedures were performed using CARTO (Biosense Webster) or EnSite (Abbott) 3D mapping systems. Both irrigated and non-irrigated catheters were used depending on operator preference and era. Contact force catheters targeted 5–40 g of force. Radiofrequency (RF) applications were delivered point by point, typically at 30 W (up to 50 W). On the posterior wall within 1 cm of the esophagus, power was limited to 20 W.

Pulmonary vein isolation (PVI) was the primary endpoint. Extra-PV ablation—including superior vena cava isolation, cavotricuspid isthmus ablation, LA roof/bottom lines, mitral isthmus line, and complex fractionated atrial electrogram (CFAE) ablation—was performed at the operator’s discretion, particularly for persistent AF or inducible AF after PVI. Sequential substrate modification was performed in a fixed order until AF termination and restoration of sinus rhythm were achieved (Figure 1).

### Ablation energy analysis

Cumulative RF energy per lesion was calculated as output (W) × duration (s), and averaged per site. In patients who underwent both PVI and mitral isthmus (MI) ablation, mean cumulative energy per lesion was compared using a paired t-test.

### CT imaging and definition of LAC

CT imaging was performed using a 320-slice multidetector scanner (Aquilion 320, Toshiba, Japan). Images were reconstructed with 3.0-mm slice thickness at pulmonary and mediastinal windows. LAC was defined as a sequential lesion with density >130 Hounsfield units (HU) and >3 pixels in size, evaluated using a dedicated workstation (Synapse Vincent, Fujifilm Co.), according to the definition of coronary artery calcification^9,10^. Calcification of the mitral annulus was excluded. For the LAC group, the time interval was measured from the first ablation to the CT date when LAC was first detected; for the non-LAC group, it was measured from the first ablation to the most recent CT date.

### Follow-up protocol

Patients were followed at 1, 3, 6, and 12 months after ablation, and annually thereafter. At each visit, 12-lead ECG and 24-hour Holter monitoring were performed at 3, 6, and 12 months, and annually thereafter, in accordance with international consensus guidelines^11^. Additional monitoring was performed if patients reported symptoms. A 3-month blanking period was applied. Recurrence of atrial tachyarrhythmia (ATA) was defined as any documented ATA beyond the blanking period.

### Clinical outcomes

The primary endpoint was a composite of cardiovascular death, HF hospitalization, ischemic stroke, or percutaneous coronary intervention. Patients without events were censored at the last clinical follow-up.

### Statistical analysis

Continuous variables were expressed as mean ± SD and compared using the Student’s t-test. Categorical variables were presented as counts (percentages) and compared using the χ² test or Fisher’s exact test, as appropriate. Predictors of LAC were identified using Cox proportional hazards models, with hazard ratios (HRs) and 95% confidence intervals (CIs) reported. Event-free survival was assessed by Kaplan–Meier analysis and log-rank testing. All analyses were performed using EZR (Saitama Medical Center, Jichi Medical University), a graphical interface for R version 4.3.3 (R Foundation for Statistical Computing, Vienna, Austria).

## Results

### Patient characteristics and ablation procedures

A total of 231 patients (mean age 66.5 ± 10.5 years, 71% men) underwent AF ablation, chest CT, and echocardiography at prespecified time intervals. Baseline characteristics are summarized in Table 1. Paroxysmal AF was present in 160 patients (69%). The median follow-up after ablation was 8.8 ± 2.6 years (until detection of LAC or last available CT).

**Table 1.**
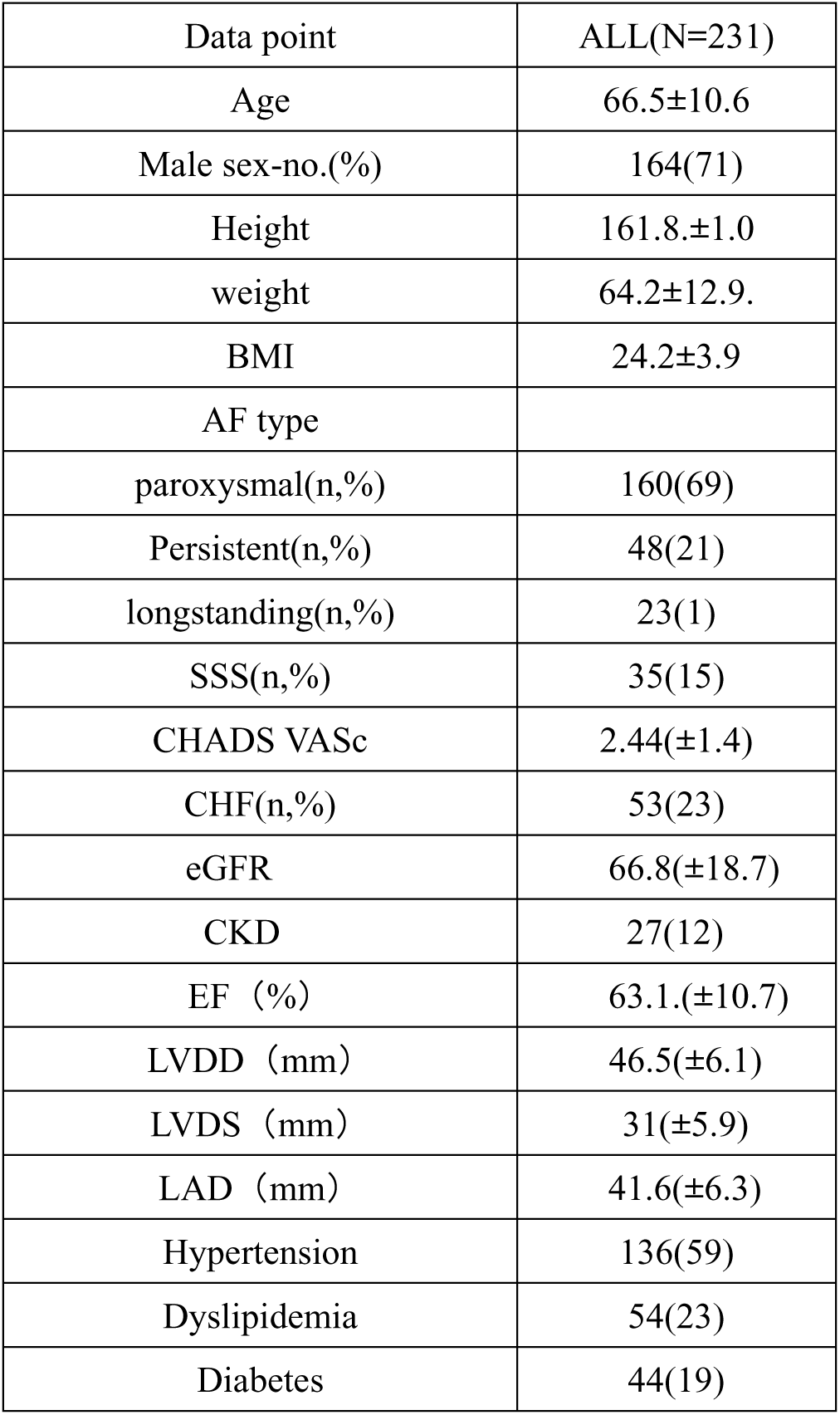
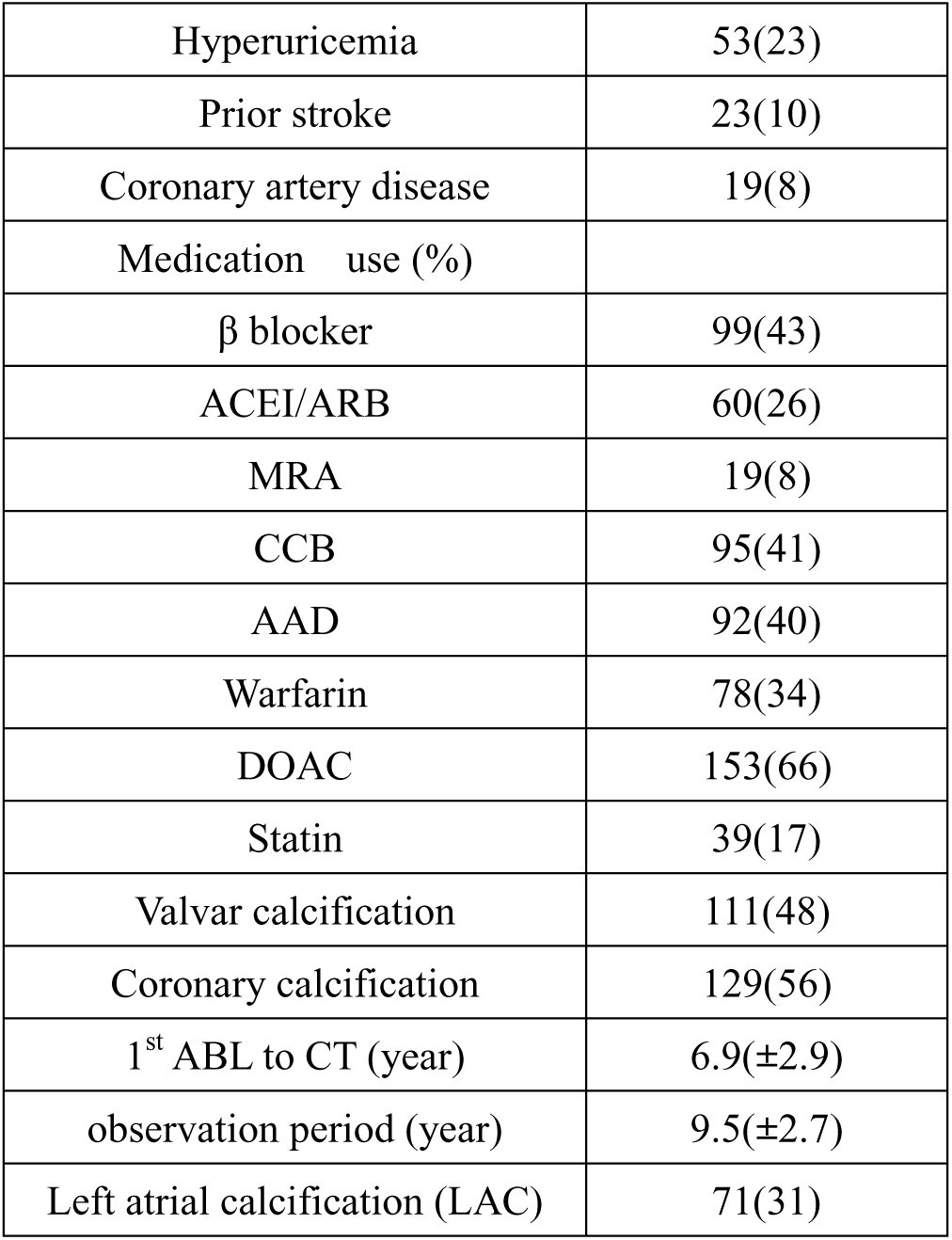
clinical characteristics of AF patients SSS=sick sinus syndrome, CHF=chronic heart failure, eGFR=estimated Glomerular Filtration Rate, CKD=Chronic Kidney Disease, EF=Ejection Fraction, ACEI/ARB=Angiotensin-Converting Enzyme Inhibitor/Angiotensin II Receptor Blocker, MRA=Mineralocorticoid Receptor Antagonist, CCB=Calcium Channel Blocker, AAD=Antiarrhythmic Drug, DOAC=Direct Oral Anticoagulant.

| Data point | ALL(N=231) |
| --- | --- |
| Age | 66.5±10.6 |
| Male sex-no.(%) | 164(71) |
| Height | 161.8.±1.0 |
| weight | 64.2±12.9. |
| BMI | 24.2±3.9 |
| AF type |  |
| paroxysmal(n,%) | 160(69) |
| Persistent(n,%) | 48(21) |
| longstanding(n,%) | 23(1) |
| SSS(n,%) | 35(15) |
| CHADS VASc | 2.44(±1.4) |
| CHF(n,%) | 53(23) |
| eGFR | 66.8(±18.7) |
| CKD | 27(12) |
| EF (%) | 63.1.(±10.7) |
| LVDD (mm) | 46.5(±6.1) |
| LVDS (mm) | 31(±5.9) |
| LAD (mm) | 41.6(±6.3) |
| Hypertension | 136(59) |
| Dyslipidemia | 54(23) |
| Diabetes | 44(19) |
| Hyperuricemia | 53(23) |
| Prior stroke | 23(10) |
| Coronary artery disease | 19(8) |
| Medication use (%) |  |
| β blocker | 99(43) |
| ACEI/ARB | 60(26) |
| MRA | 19(8) |
| CCB | 95(41) |
| AAD | 92(40) |
| Warfarin | 78(34) |
| DOAC | 153(66) |
| Statin | 39(17) |
| Valvar calcification | 111(48) |
| Coronary calcification | 129(56) |
| 1<sup>st</sup> ABL to CT (year) | 6.9(±2.9) |
| observation period (year) | 9.5(±2.7) |
| Left atrial calcification (LAC) | 71(31) |

Valvular calcification and coronary calcification were present in 48% and 56% of patients, respectively. The mean time from first ablation to CT was 6.9 ± 2.9 years, while the mean clinical follow-up was 9.5 ± 2.7 years. Overall, 31% (71/231) of patients developed LAC.

Details of ablation procedures are shown in Table 2. PVI was performed in all patients. Multiple ablations were required in 47%, and 36% underwent extra-PV ablation.

**Table 2.** Patient characteristics related to the ablation procedure.

|  |  |
| --- | --- |
| No. ablation procedures |  |
| 1 | 123(53) |
| 2 | 66(29) |
| 3 or more | 22(18) |
| Contact force catheter | 112(48) |
| PVI | 231(100) |
| Extra PV LA ablation | 84(36) |
| Linear ablation | 60(26) |
| Roof line ablation | 52(23) |
| Bottom line ablation | 43(19) |
| Mitral Isthmus ablation | 42(18) |

PVI=Pulmonary Vein Isolation, PV=Pulmonary Vein, LA=Left Atrium

### Characteristics of patients with LAC

Baseline differences between the LAC and non-LAC groups are shown in Table 3. Compared with the non-LAC group, patients with LAC were younger (64.7 ± 8.3 vs. 67.2 ± 11.3 years, p<0.01), more frequently had longstanding AF (21% vs. 5%, p<0.01), and exhibited larger LA diameters (44.7 ± 6.1 vs. 40.0 ± 6.0 mm, p<0.01). The LAC group also more often required multiple ablations and extra-PV ablation.

**Table 3.**
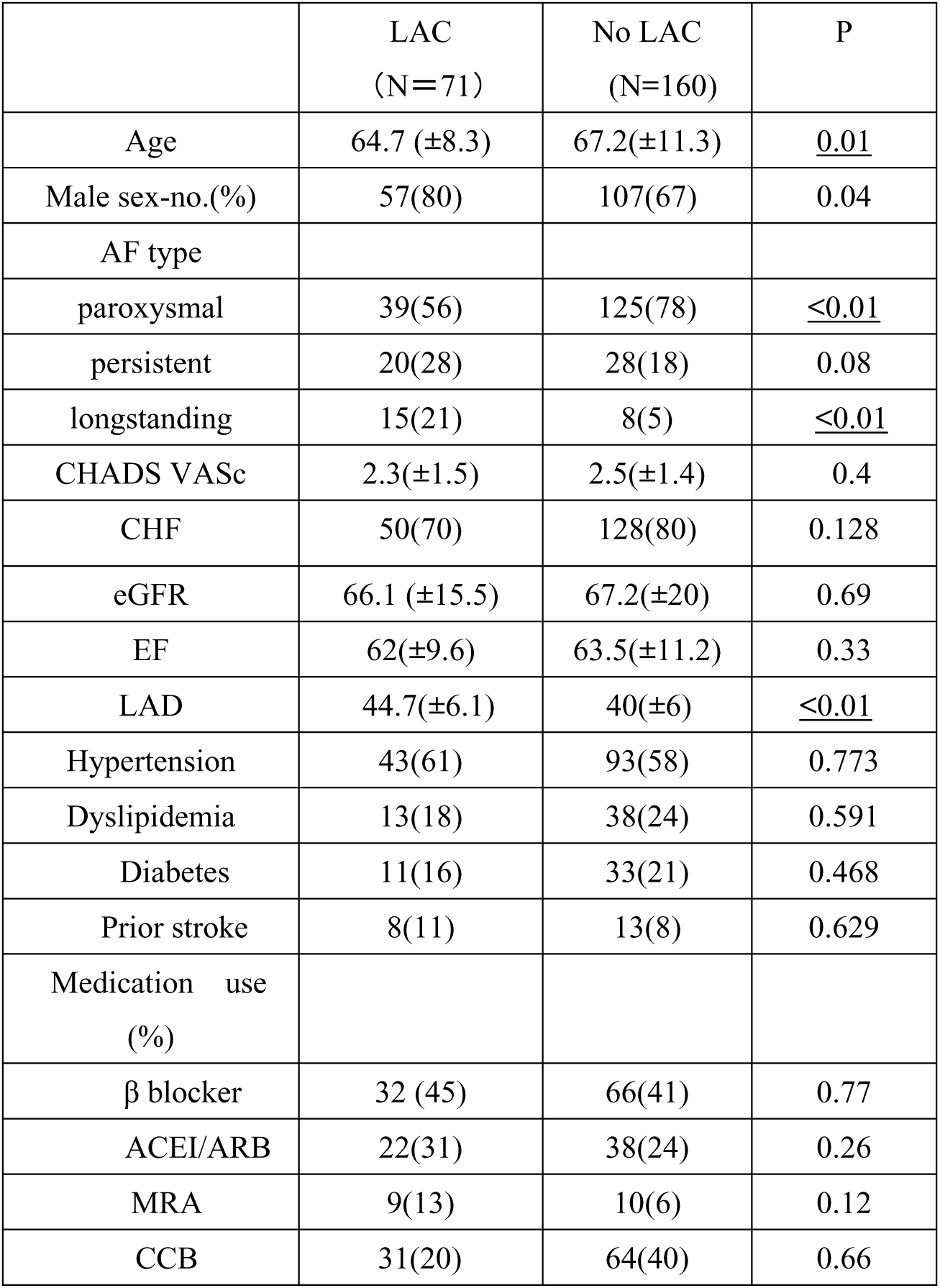
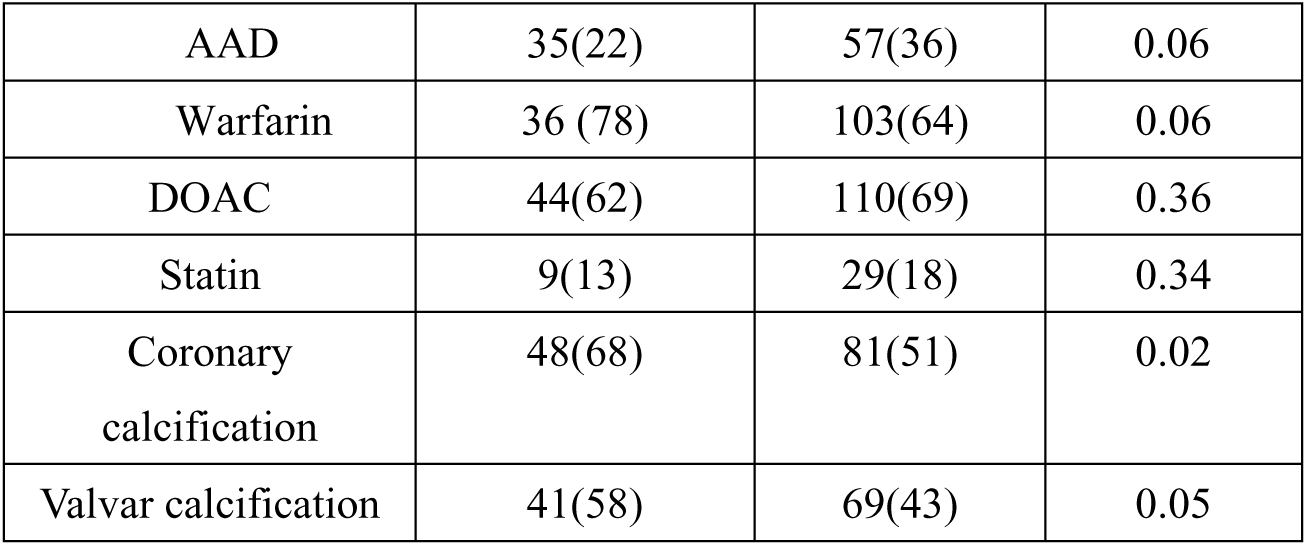
Comparison in clinical characteristics between LAC group and non-LAC group.

**Table 4.** Comparison in Ablation data between LAC group and non-LAC group.

| Procedure | LAC | Non-LAC |  |
| --- | --- | --- | --- |
| PVI | 71(100) | 160(100) | 1 |
| Roof | 36(51) | 30(19) | <u><0.01</u> |
| Bottom | 35(49) | 28(18) | <u><0.01</u> |
| Posterior Wall | 23(33) | 20(13) | <u><0.01</u> |
| MI | 38(54) | 18(11) | <u><0.01</u> |
| LAA | 29(41) | 14(9) | <u><0.01</u> |
| septum | 32(45) | 16(10) | <u><0.01</u> |
| Defragmentation | 28(39) | 18(11) | <u><0.01</u> |
| Two or more ABL | 55(78) | 60(38) | <u><0.01</u> |
| ABL other than PV | 49(69) | 35(22) | <u><0.01</u> |

MI=Mitral Isthmus, LAA=Left Atrial Appendage

The interval from ablation to detection of LAC was 7.3 ± 3.1 years, most frequently between 6 and 8 years (minimum 2.7 years) (Figure 3). The distribution of calcification sites is presented in Figure 4: the lateral mitral annulus (20%), anterior RSPV (11%), and posterior RIPV (10%) were the most common locations. The proportion of calcified sites relative to ablation at the corresponding location is shown in Table 5, with the highest rate at the lateral annulus (45%). Total ablation time and energy per point were greater at the mitral isthmus in patients with LAC (Figure 5).

**Figure 3.**
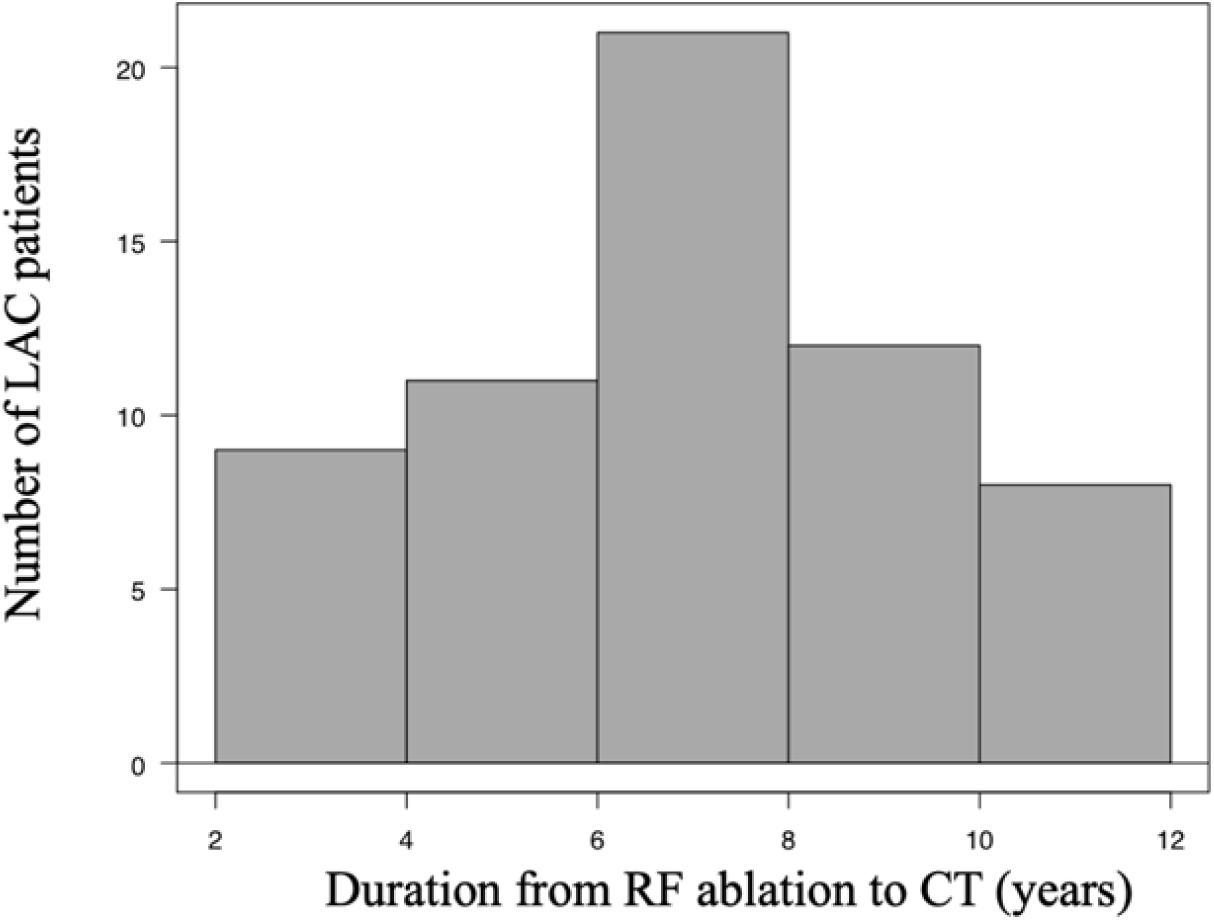
Time from ablation to finding LAC on CT.

**Figure 4.**
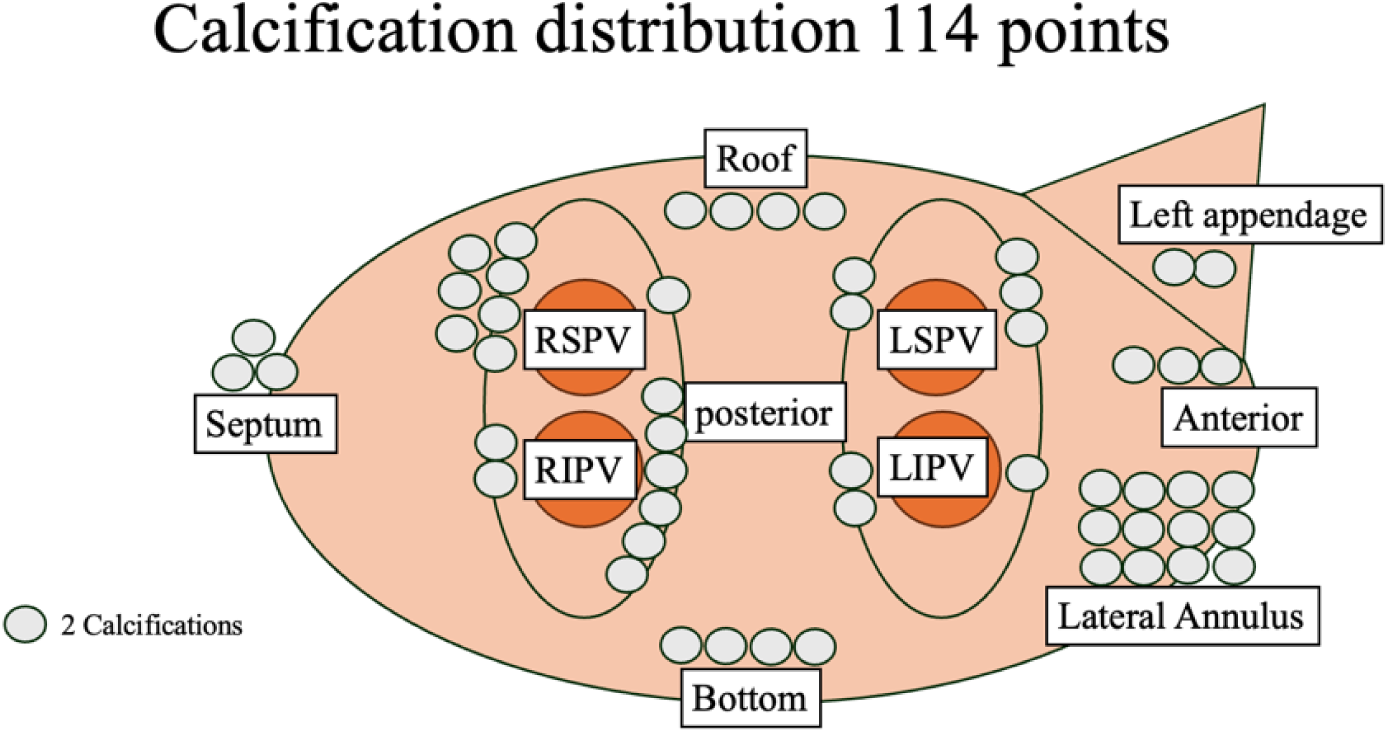
Calcification distribution. Total 114 points.

**Figure 5.**
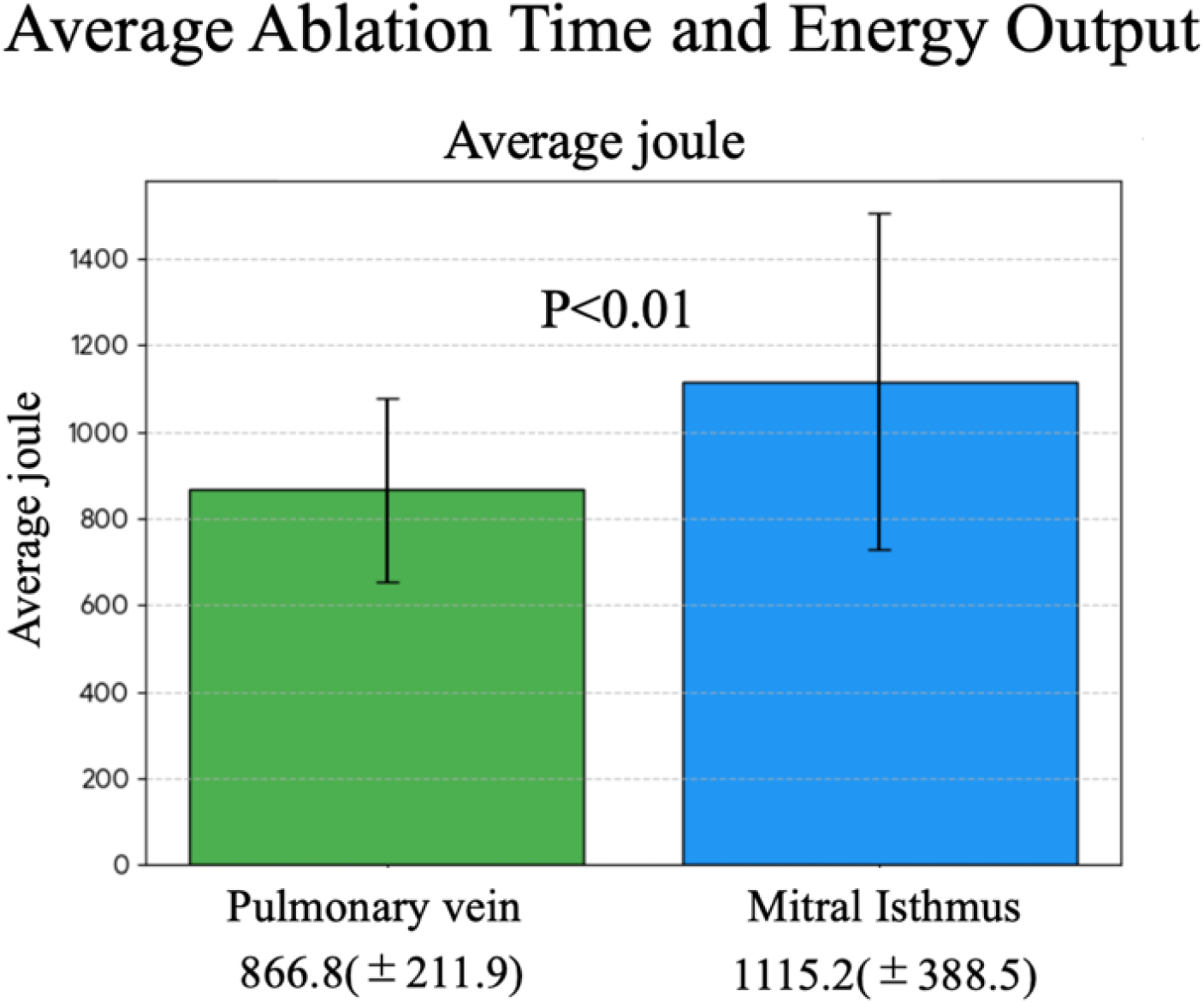
Pulmonary Vein vs. Mitral Isthmus Ablation: Comparison of Ablation Energy (Joules)

**Table 5.** Ratio of total number of calcifications and people ablated (%)

|  | LAC |  | LAC |  | LAC |
| --- | --- | --- | --- | --- | --- |
| Lateral annulus | 25/56(45%) | LSPV anterior | 7/231(3%) | RSPV anterior | 14/231 (6%) |
| Roof | 9/66(14%) | LSPV posterior | 4/231(2%) | RSPV posterior | 3/231 (1%) |
| Bottom | 8/63(13%) | LIPV anterior | 8/231(4%) | RIPV anterior | 5/231 (2%) |
| LAA | 3/41(7%) | LIPV posterior | 4/231(2%) | RIPV posterior | 12/231 (5%) |
|  |  | LA Septum | 6/48(13%) | Anterior Wall | 6/41(15%) |

In multivariate Cox regression (Table 6), independent predictors of LAC formation included:

- non-paroxysmal AF (HR 1.7, 95% CI 1.0–2.8, p=0.04),
- LA diameter >50 mm (HR 2.7, 95% CI 1.3–5.5, p<0.01), and
- extra-PV ablation (HR 2.3, 95% CI 1.3–4.2, p=0.04).

**Table 6.**
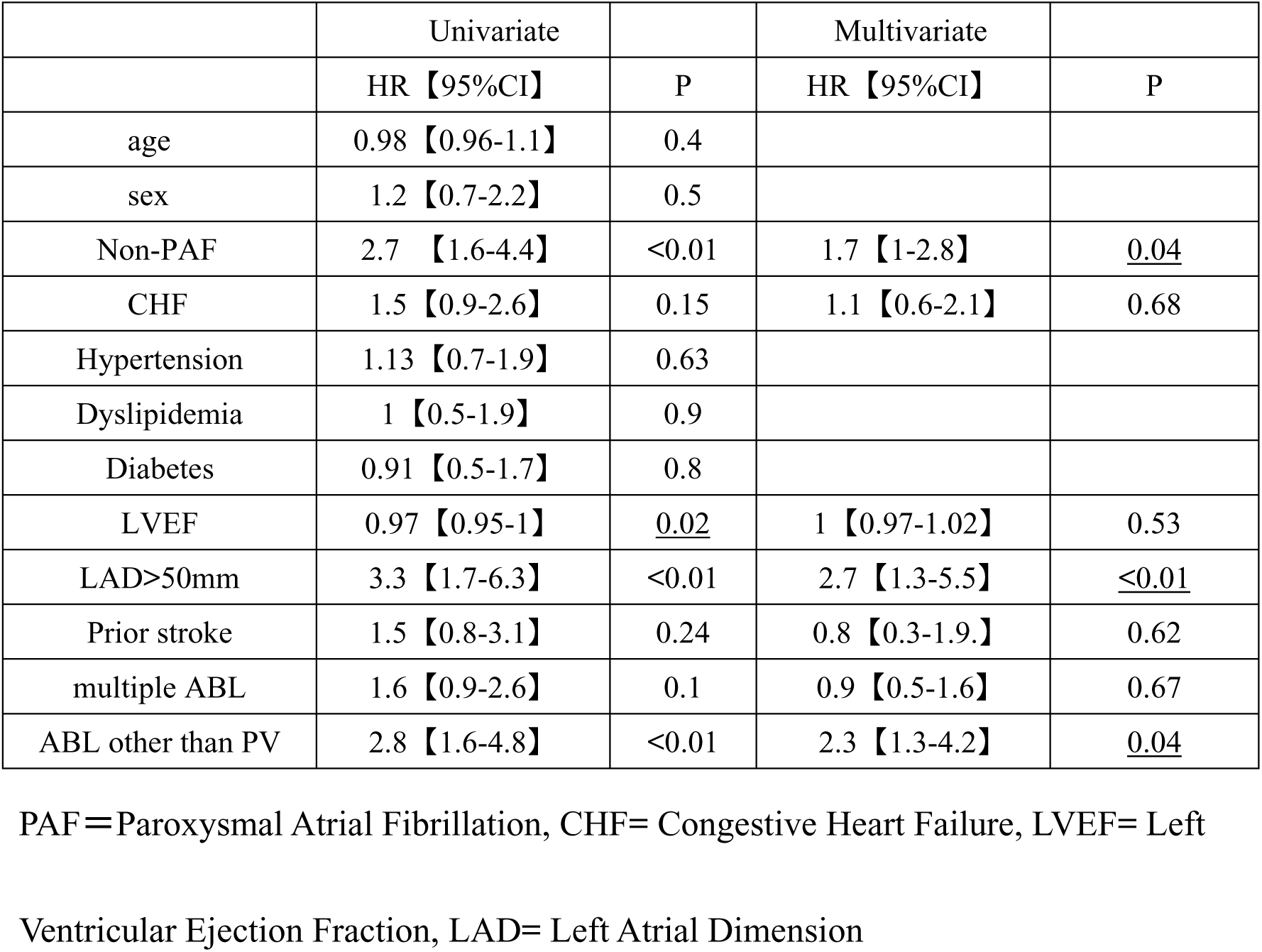
Cox Hazards Analysis for the Predictors of LAC After Ablation.

### Prognosis of patients with LAC

Kaplan–Meier analysis demonstrated a significantly higher incidence of HF hospitalization in the LAC group compared with the non-LAC group (Figure 6, p=0.02). In contrast, there was no difference in the incidence of stroke (Figure 7, p=0.50). The composite endpoint of HF hospitalization, stroke, or coronary intervention was more frequent in the LAC group (Figure 8, p=0.05).

**Figure 6.**
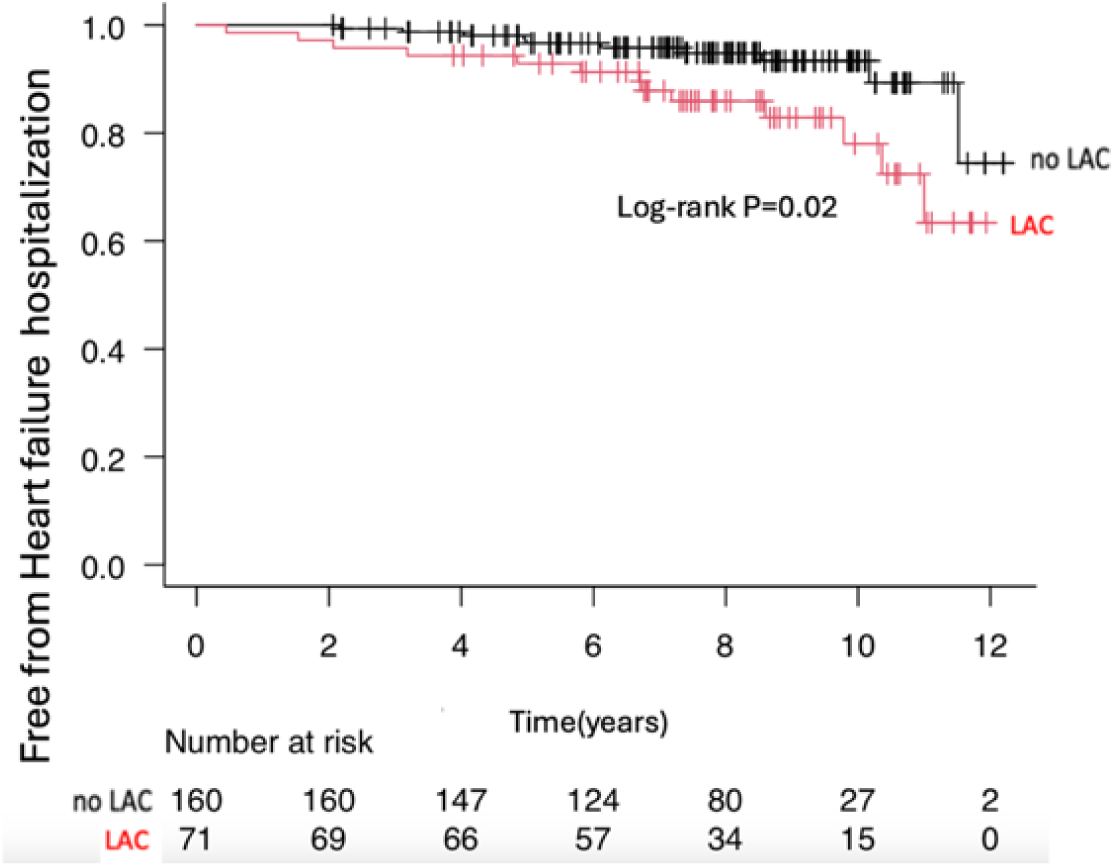
Differences in survival curves for development of heart failure in the LAC and non-LAC groups.

**Figure 7.**
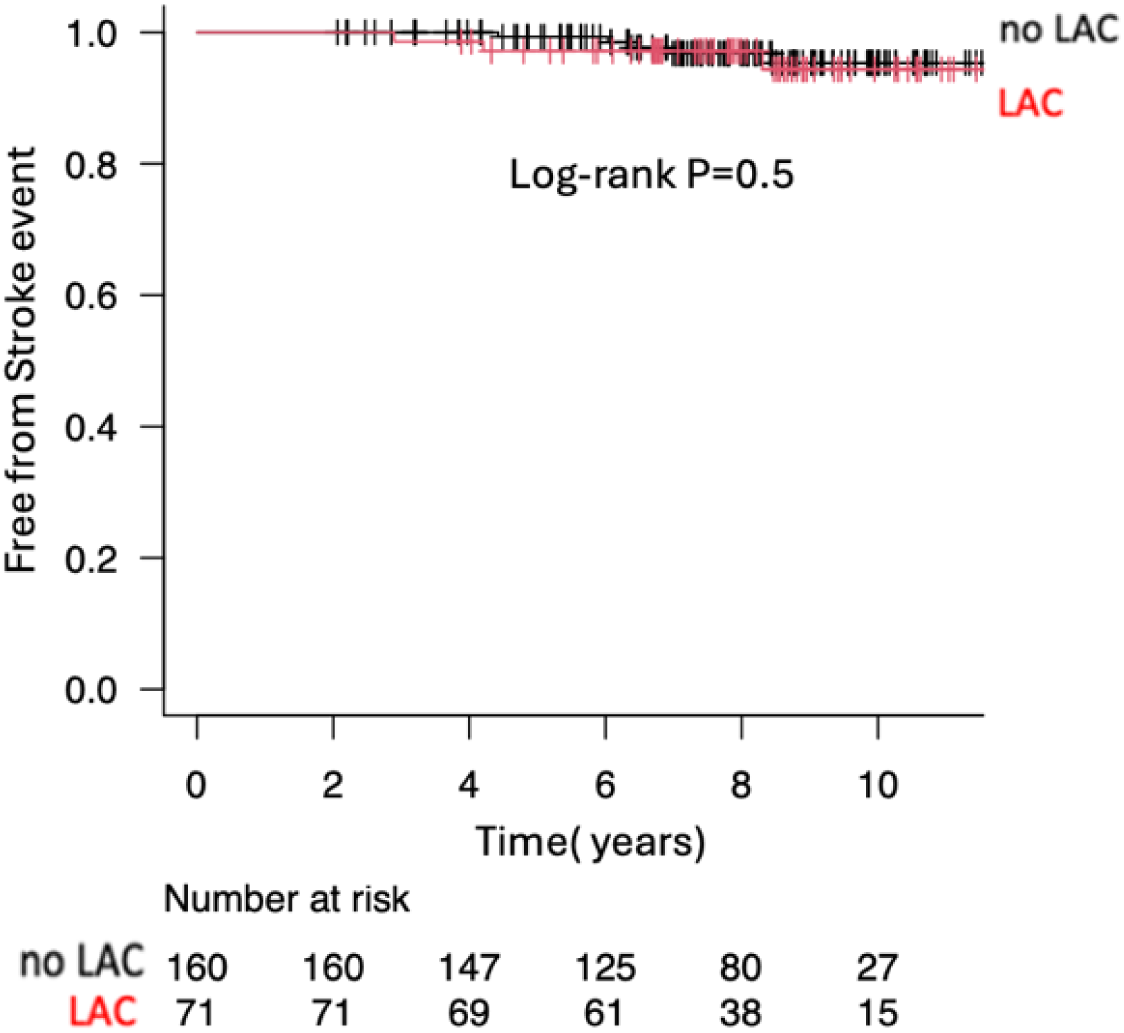
Differences in survival curves for development of stroke in the LAC and non-LAC groups.

**Figure 8.**
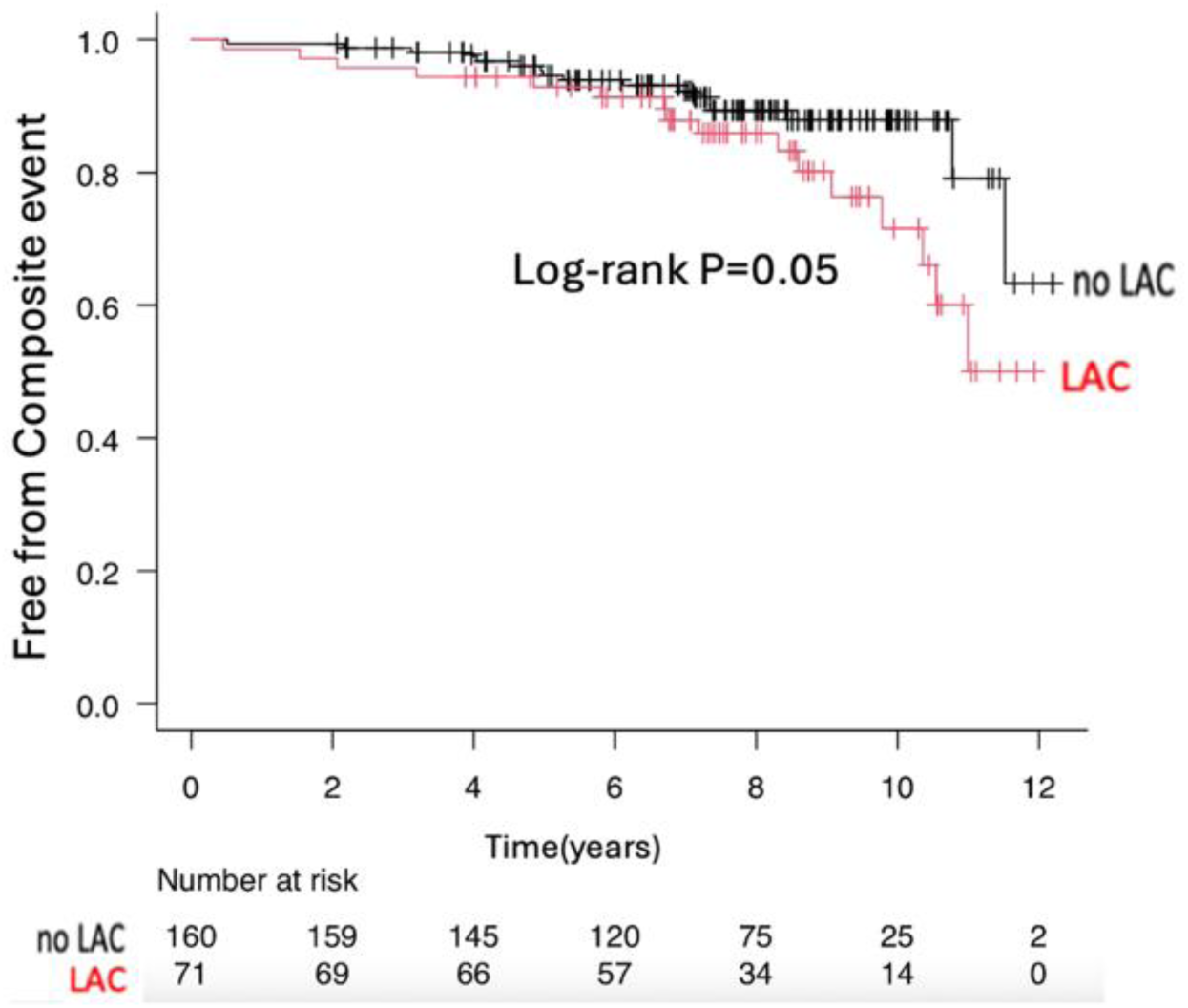
Differences in survival curves for composite endpoints in the LAC and non-LAC groups.

## Discussion

### Main findings

In this study, we demonstrated that (i) non-paroxysmal AF, LA diameter >50 mm, and extra-PV ablation were independent predictors of LAC formation after ablation, (ii) calcification occurred not only at pulmonary vein sites but was most frequent at the lateral mitral annulus (MI line), and (iii) patients with LAC had significantly higher rates of HF hospitalization compared with those without LAC. To our knowledge, this represents one of the largest and longest observational studies investigating the predictors and clinical consequences of LAC after AF ablation.

### LAC after RF ablation

LAC has traditionally been reported in association with rheumatic heart disease, mitral valve disease, and organized thrombi^3–5^. More recently, several studies have described LAC after AF ablation^6^. Yakabe et al. reported that post-ablation LAC was associated with increased cardiovascular events^12^. Our study extends these findings by providing longer follow-up (mean 9.5 years vs. 6 years in Yakabe et al.), analyzing procedural factors, and identifying specific ablation sites as risk modifiers for LAC development. These results suggest that ablation itself may serve as an etiology of acquired LA calcification in the chronic phase.

### Predictors of LAC formation

Independent predictors of LAC included non-paroxysmal AF, larger LA diameter, and extra-PV ablation. Patients with advanced atrial remodeling (non-PAF and LA enlargement) typically require more extensive ablation, which may increase tissue injury and subsequent calcification. Similar to dystrophic cardiac calcinosis described in animal models^13^, chronic injury and repair processes may promote calcification in atrial tissue.

The identification of extra-PV ablation as a predictor is noteworthy. In particular, ablation at the mitral isthmus was strongly associated with subsequent calcification. This site is technically challenging due to thick atrial myocardium, epicardial connections, and convective cooling from coronary sinus flow, often necessitating higher cumulative RF energy^14,15^. In our cohort, greater energy delivery at the MI line was correlated with subsequent calcification, suggesting that excessive thermal injury and inflammation may trigger fibrosis and calcium deposition.

### Sites of calcification formation

Prior studies suggested that calcification after AF ablation is most common in the pulmonary veins^6^. In contrast, our data demonstrated a higher relative incidence at the mitral isthmus and lateral annulus. The frequent occurrence at the MI line, potentially exacerbated by the high-energy ablation targeting the pulmonary veins in this study, suggests that RF-induced injury may be the predominant mechanism of calcification. These findings highlight the importance of procedural strategy in minimizing long-term atrial damage. Additional procedures such as ablation in the coronary sinus and ethanol injection into the Marshall vein are also options for the purpose of reducing energy at high frequency from the endocardium^16,17^.

### Clinical implications of LAC

Patients with LAC experienced higher rates of HF hospitalization during long-term follow-up. This aligns with case-based observations suggesting that extensive atrial calcification may contribute to impaired compliance, pulmonary hypertension, and HF progression^18^. Although stroke incidence did not differ between groups, isolated reports of mobile LAC fragments causing embolic events underscore the need for continued vigilance. Importantly, calcification was observed as early as 2.7 years after ablation, suggesting that preprocedural imaging during repeat ablation should include careful evaluation for LAC.

### Limitations

This was a retrospective, single-center study, and the sample size of patients undergoing CT was relatively modest compared with the overall AF ablation population.

Nevertheless, the long follow-up duration and detailed procedural analysis strengthen the validity of our findings. Prospective multicenter studies are warranted to confirm these results and to clarify the mechanistic links between RF ablation, atrial injury, and calcification.

## Conclusion

LAC after AF ablation is not a rare phenomenon and is independently predicted by non-paroxysmal AF, LA enlargement, and extra-PV ablation. Calcification occurs not only in the pulmonary veins but frequently at the mitral isthmus, likely related to greater energy delivery at this site. Importantly, LAC is associated with higher rates of HF hospitalization, underscoring its significance as a chronic complication of AF ablation and a potential imaging marker for long-term risk stratification.

## Abbreviations

LAC: left atrial calcification
CFAE: Complex fractionated atrial electrograms
PV: pulmonary vein
PVI: pulmonary vein isolation
HF: heart failure
LA: Left atrium
CT: Computed Tomography
MI: Mitral isthmus

## Data Availability

All data generated or analyzed during this study are included in this published article and its supplementary information files.

## Acknowledgment

The authors have no acknowledgments to declare.

